# Predicting Cardiopulmonary Exercise Testing Performance in Patients Undergoing Transthoracic Echocardiography - An AI Based, Multimodal Model

**DOI:** 10.1101/2025.07.05.25330921

**Authors:** Shudhanshu Alishetti, Weishen Pan, Ashley N. Beecy, Zhenzhen Liu, Albert Gong, Zhe Huang, Kevin J. Clerkin, Rochelle L. Goldsmith, David T. Majure, Chris Kelsey, David vanMaanan, Jeffrey Ruhl, Naomi Tesfuzigta, Erica Lancet, Deepa Kumaraiah, Gabriel Sayer, Deborah Estrin, Kilian Weinberger, Volodymyr Kuleshov, Fei Wang, Nir Uriel

**Author notes:** **Address for correspondence:** Shudhanshu Alishetti, MD 506 Sixth St, 2nd Floor, Brooklyn, NY 10021. Co-first authors (Shudhanshu Alishetti, Weishen Pan). These authors contributed equally. The institution where the work was performed: NewYork-Presbyterian/Columbia University Irving Medical Center, NewYork-Presbyterian/Weill Cornell Medical Center, NewYork-Presbyterian/Brooklyn Methodist Hospital NewYork-Presbyterian/Queens Hospital.

## Abstract

**Background and Aims:** Transthoracic echocardiography (TTE) is a widely available tool for diagnosing and managing heart failure but has limited predictive value for survival. Cardiopulmonary exercise test (CPET) performance strongly correlates with survival in heart failure patients but is less accessible. We sought to develop an artificial intelligence (AI) algorithm using TTE and electronic medical records to predict CPET peak oxygen consumption (peak VO_2_) ≤ 14 mL/kg/min.

**Methods:** An AI model was trained to predict peak VO_2_ ≤ 14 mL/kg/min from TTE images, structured TTE reports, demographics, medications, labs, and vitals. The training set included patients with a TTE within 6 months of a CPET. Performance was retrospectively tested in a held-out group from the development cohort and an external validation cohort.

**Results:** 1,127 CPET studies paired with concomitant TTE were identified. The best performance was achieved by using all components (TTE images, all structured clinical data). The model performed well at predicting a peak VO_2_ ≤ 14 mL/kg/min, with an AUROC of 0.84 (development cohort) and 0.80 (external validation cohort). It performed consistently well using higher (≤ 18 mL/kg/min) and lower (≤ 12 mL/kg/min) cut-offs.

**Conclusions:** This multimodal AI model effectively categorized patients into low and high risk predicted peak VO_2_, demonstrating the potential to identify previously unrecognized patients in need of advanced heart failure therapies where CPET is not available.

## Introduction

Heart failure (HF) impacts nearly 7 million in the United States, with that number expected to increase to more than 10 million by 2040(1). HF is a progressive, fatal condition that is associated with a median loss of 15 years of life. It has the potential to impact all, agnostic to age, ethnicity, or location; however, some groups bear a greater burden. Patients in rural areas demonstrate higher HF mortality for both younger and older age groups, with mortality rates rising steadily since 2012. Despite improvements in medical therapy, HF related mortality is on the rise. One potential reason is inadequate awareness and recognition of patients with advanced HF. This subset of patients, an estimated 200,000 Americans, have survival worse than most cancers(1,2). Advanced therapies (heart transplantation and left ventricular assist devices) can alter that trajectory, improving survival and quality of life. Without advanced therapies, patients with advanced heart failure have a one-year survival of 25-50%(3,4). Yet only about 6,000 patients receive advanced HF therapies annually; the other 194,000 do not.

Echocardiography plays a significant role in the diagnosis and management of patients with HF. It is ubiquitously available and provides an assessment of cardiac structure and function. However, it has had limited value in predicting survival in HF. Secondary analyses of the Val-HeFT and CHARM studies(5,6) demonstrated worse survival with decreasing EF. However, the SOLVD trial saw worse survival in patients with symptomatic HF compared with asymptomatic HF, and interestingly did not see a significant difference in EF between patients with asymptomatic HF and those with moderate HF symptoms(7,8). More recently a study of almost 1,000 patients who met the European Society of Cardiology definition of advanced heart failure found that ejection fraction was not associated with hospitalization or mortality(3). Thus, while an important test, it cannot faithfully identify patients who have progressed from symptomatic HF (Stage C) to advanced HF (Stage D) where advanced therapies are necessary.

Functional capacity has demonstrated a strong association with outcomes in HF and identification of advanced HF. Across numerous studies, patients with New York Heart Association (NYHA) functional class I symptoms had a 1 year mortality around 5%, rising to 15% for those with NYHA class II or III symptoms, and 44% for NYHA class IV(7–9). Cardiopulmonary exercise testing (CPET) allows for further delineation of functional capacity beyond the NYHA functional classes. A seminal study by Mancini et al. found that peak VO_2_, an objective measure of exercise capacity, predicted survival and differentiated between when patients needed heart transplantation and when transplantation could be safely deferred(10). Performance on these tests also correlates with HF prognosis(11,12). However, CPETs are far less frequently administered than TTE. This is in part because of a limited number of facilities that perform CPETs as they require an exercise testing laboratory equipped with a metabolic cart as well as a medical professional capable of interpreting the test.

Artificial intelligence (AI) applied to echocardiography has demonstrated the ability to quantify ejection fraction(13–15), quantitate cardiac structures(16), assess mitral regurgitation(17), detect various disease states, and predict outcomes(18–20). While such models aim to obtain information that trained echocardiography readers can gather from a study, AI techniques may allow for the extraction of information from echocardiograms that cannot currently be obtained using usual techniques, such as exercise capacity. In this study we sought to develop and evaluate an AI algorithm utilizing multi-modal architecture incorporating echocardiography data and demographic data readily available in an electronic medical record system to predict peak VO_2_.

## Methods

### Ethics approval

This study was approved by the institutional review boards at Weill Cornell Medicine and Columbia University Irving Medical Center. A waiver for informed consent was obtained.

### Study Cohort

This study leveraged a database from different sites of NewYork Presbyterian/Columbia University Irving Medical Center (CUMC), NewYork Presbyterian/Weill Cornell Medical Center (WCM), NewYork Presbyterian/Brooklyn Methodist Hospital, and NewYork Presbyterian Queens Hospital. The database includes data from cardiopulmonary exercise tests (CPET), transthoracic echocardiograms (TTE), and clinical data from the electronic health record (EHR) outlined below. Considering the sample sizes of the sites, data from CUMC were used to construct the training samples, while the samples from all the other sites were used for external validation.

### Data Curation and Preprocessing

The study cohort included adult (age ≥ 18 years) patients at CUMC who underwent a CPET with adequate effort (respiratory exchange ratio (RER) ≥ 1.05) within six months of a transthoracic echocardiogram (TTE).

#### Development Cohort Samples

Since 2017 3,872 cardiopulmonary exercise testing studies have been conducted for adult patients at CUMC within 6 months of a TTE. Of those, 1,000 met the specific inclusion criteria and were used to train the model. Among the patients excluded, 2,454 patients were excluded because TTE images and videos were not available, 32 were excluded because TTEs were in the pediatric format, and 12 were excluded because no available TTE videos from the standard parasternal long axis (PLAX), apical four chamber (A4C) view types were in the corresponding echo studies, and 374 were excluded because of a sub-maximal CPET (respiratory exchange ratio<1.05) (Supplemental Figure 1).

#### External Validation Samples

An external validation set of 127 CPET studies which met inclusion criteria was compiled consisting of patients who had cardiopulmonary exercise testing completed at WCM, NewYork Presbyterian/Brooklyn Methodist Hospital, and NewYork Presbyterian Queens Hospital (Supplemental Figure 1).

To construct the input, we first preprocessed the data by extracting the structured features from the electronic health records (EHRs) and identifying the types of videos and images from an TTE as follows:

#### Structured Feature Extraction and Preprocessing

Different categories of variables were considered to construct the structured feature vector to predict peak VO_2_, including demographics, concomitant medications, laboratory tests, vital signs and structural TTE measurement and findings. These variables were determined by clinical experts. The full list of the variables is presented in the Supplementary Table 1. And the preprocess procedures for different kinds of variables are illustrated as follows:

##### Demographic variables

Demographic variables included age at the CPET date, sex, and race.

##### Concomitant medications

Medications were grouped into pharmaceutical subclasses based on the anatomical therapeutic chemical (ATC) codes. Each medication feature denotes whether the medications from its respective group were used in the observation window.

##### Laboratory tests and vital signs

Laboratory tests and vital signs were identified from structured data using Logical Observations Identifier Names and Codes (LOINC) within the observation period. For each test, the value closest to the CPET date was utilized in the presence of multiple measurements.

##### TTE measurement and findings

The features of measurement and findings were extracted from the report of echo study closest to the target CPET.

After constructing the features, the missing values were imputed as: (1) mode for categorical and binary features; and (2) mean for continuous features. The mode and mean values for imputation were calculated based on the training set of the development data (see Model Training, Development, and Validation for data splitting). In order to eliminate the effects of value magnitude, all variables were scaled according to the z-score.

#### TTE Video/Image Identification and Preprocessing

For TTE data, we classified the subtypes of TTE videos without color Doppler, M-mode images, and color Doppler images, respectively. The preprocess procedures for different kinds of TTE imaging data are illustrated as follows.

##### TTE videos without color Doppler

Specifically, we utilized a convolutional neural network (CNN) to identify the TTE videos as one of the standard echocardiographic views. In particular, we utilized the pre-trained model in the previous study to classify the TTE videos, which is publicly available and widely used(13). With the pre-trained image-based classifier, we first fed each frame sub-sampled from a video to the classifier and averaged the resulting probability distributions to obtain the prediction result of the video. Finally, we selected the TTE videos with the standard parasternal long axis (PLAX), apical four chamber (A4C) views. After identifying and selecting the TTE videos, we further preprocessed the selected videos by cropping and masking them to remove the text and all other information outside the scanning sector. The resulting videos were further subsampled by one in every two frames and resized to 112 × 112-pixel video.

##### M-mode images

We manually labeled and selected the images based on the focused regions including the aortic valve (AO), mitral valve (MV) and left ventricle (LV). The selected M-mode images were further cropped to only keep the waveform region of which the coordinates were annotated in the Digital Imaging and Communications in Medicine (DICOM) files. The resulting images were resized to 224 × 224-pixel images.

##### Color Doppler images

We first divided them into tissue Doppler images, Doppler images with pulsed wave (PW) and Doppler Images with continuous wave (CW) based on the tag stored in the DICOM files. Then we trained CNNs to identify subtypes of these color Doppler images based on the focused regions and measurements with the labeled data from the external WCM cohort with available annotations and apply it to the data from the CUMC cohort. The selected Doppler images were further cropped to only keep the waveform region and remove the artifacts in the waveform region. The resulting images were resized to 224 × 224-pixel images. Supplementary Table 2 lists the types of TTE videos and images used in our model.

### Model Structure

After extracting the structured features and identifying the TTE videos and images, we fed each into two separate branches to calculate the respective prediction scores. Finally, we assembled the prediction scores obtained from the structured features and TTE data to get the final prediction, as illustrated in Figure 1.

**Figure 1.**
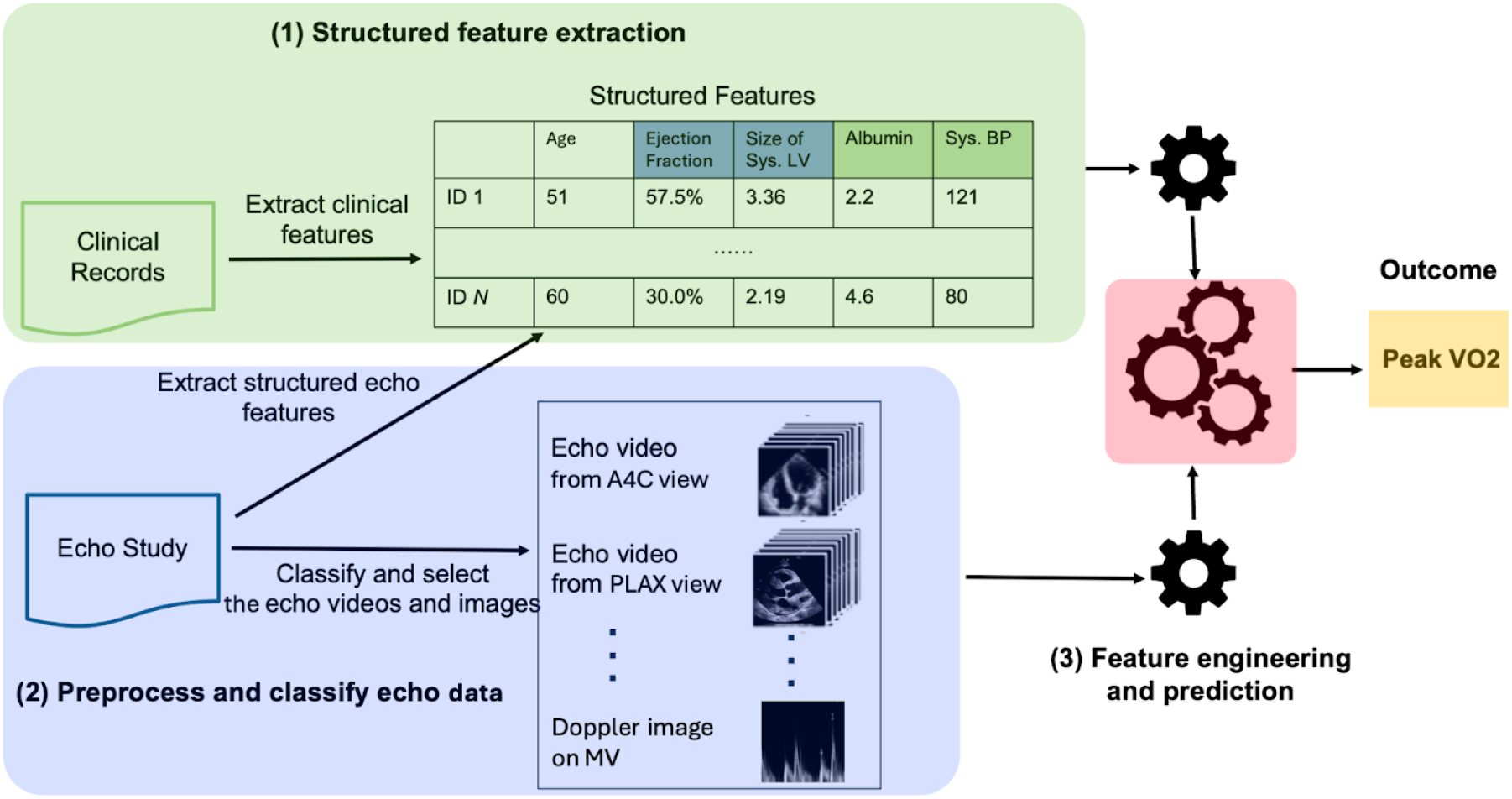
The workflow of the method.

In the branch with the structured features, we tested 4 popular models including the logistic regression, multilayer perceptron random forest, and extreme gradient boosting models. The branch of the model with the TTE data is illustrated in Figure 1(b). Since multiple TTE videos and images existed in each TTE study with arbitrary numbers from each subtype, we first fed the videos/images into category-specific encoders based on their categories (TTE videos without color Doppler, m-mode images, or color Doppler images) to extract the features. Then the extracted features were input into subtype-specific regressors based on their subtypes (e.g., from A4C or PLAX for TTE videos without color Doppler) to obtain the predicted peak VO_2_. We repeated the above procedure for all identified videos and images from the same study and then averaged the obtained predicted values as the final prediction for the outcome.

In particular, we implemented the video encoder as a 3D residual convolutional neural network (21), which is the structure used in Echonet (14). And we implemented the image encoder for m-mode and Doppler images as a 2D residual convolutional neural network.

### Video Encoder Pretraining

We first pretrained the video encoder on the publicly available Echonet-Dynamic dataset, and then fine-tuned on our CPET dataset. The Echonet-Dynamic dataset consists of 10,030 grayscale A4C videos with ejection fraction labels. For the pretraining, we followed the standard training procedure of Echonet and used an 18-layer (2+1)D ResNet (22) for ejection fraction prediction.

### Model Training, Development, and Validation

We performed a 64%-16%-20% train-validation-test split, divided by patients, within the development cohort. To train the proposed model, we first generated video-label/image-label pairs by mapping the peak VO_2_ label to each identified video and image from the linked TTE study. Then we trained the model by minimizing the mean square error on the training video-label/image-label pairs. During testing, we applied the trained model to each video/image from the same TTE study and then ensembled their results together. The training subset was used to train the model, with the hyperparameters tuned with the validation subset. Then the model was evaluated on the test subset of the development cohort, as well as the external validation cohort. We conducted the experiments with multiple randomly train-validation-test splits and reported the averaged performance.

We evaluated the performance of the model to predict a peak VO_2_ using a cut-off of ≤14 ml/kg/min. Additionally, we evaluated the model’s performance with the following tasks: (1) binary classification of peak VO_2_ using different cut-offs (≤12 mL/kg/min and ≤18 mL/kg/min); (2) directly regress the peak VO_2_ value.

## Results

### Data Curation and Preprocessing

The study cohort included 1,127 total CPET studies in the development (n=1,000) and external validation cohorts (n=127). The characteristics of the two cohorts are shown in Table 1. The two populations were similar with respect to age (mean 56.8±15.3 years vs. 56.0±15.8 years) and sex (35.8% female vs 35.4% female). The racial composition differed between the two cohorts with a greater proportion of White (47.8% vs. 40.2%) and other/declined (36.2% vs. 22.0%) patients and fewer Asian (2.5% vs. 18.1%) and Black patients (13.5% vs. 19.7%) in the development cohort. Baseline echocardiographic parameters were similar between the groups with EF (37.2%±16.7 vs. 39.9%±17.0), LV end diastolic diameter, and wall thickness having no clinically meaningful differences. Lastly the baseline peak VO_2_ measurements were higher in the development cohort than the external validation cohort (20.1±7.7 mL/kg/min vs. 18.1±8.6 mL/kg/min).

**Table 1.** Characteristics of the Development (Columbia) and External Validation (NYP-Brooklyn Methodist, NYP Queens, and Cornell) cohorts.

|  | Development<br>(n = 1,000) | External Validation<br>(n = 127) |
| --- | --- | --- |
| **Age**, years, mean ± SD | 56.75±15.30 | 56.02±15.86 |
| **Sex**, n (%) |  |  |
| Male | 632 (63.2%) | 82 (64.6%) |
| Female | 358 (35.8%) | 45 (35.4%) |
| **Race**, n (%) |  |  |
| White | 478 (47.8%) | 51 (40.2%) |
| Black | 135 (13.5%) | 25 (19.7%) |
| Asian | 25 (2.5%) | 23 (18.1%) |
| Other | 362 (36.2%) | 28 (22.0%) |
| **Echocardiographic Measures**, mean ± SD | | |
| LV ejection fraction, % | 37.19±16.74 | 39.87±16.95 |
| LV end diastolic diameter, mm | 5.62±1.37 | 5.55±1.18 |
| LV end systolic diameter, mm | 4.81±1.69 | 4.56±1.47 |
| IVS wall thickness, mm | 0.92±0.23 | 0.95±0.25 |
| **CPET Measures** |  |  |
| Peak VO<sub>2</sub>, ml/kg/min | 20.08±7.71 | 18.09±8.59 |
| **Diagnosis**, n (%) |  |  |
| Heart failure | 458 (45.8%) | 80 (63.0%) |
| -HFrEF | 401 (40.1%) | 71 (56.0%) |
| - HFpEF | 33 (3.3%) | 8 (6.3%) |
| -Right heart failure | 24 (2.4%) | 1 (0.8%) |
| Heart failure causes | 144 (14.4%) | 34 (26.7%) |
| -Ischemic cardiomyopathy | 90 (9.0%) | 15 (11.8%) |
| -Hypertensive cardiomyopathy | 54 (5.4%) | 8 (6.3%) |
| -Dilated cardiomyopathy | 60 (6.0%) | 11 (8.7%) |
| -HCM | 118 (11.8%) | 14 (11.0%) |
| -Sarcoid | 13 (1.3%) | 0 (0.0%) |
| Valve Disease | 188 (18.8%) | 21 (16.5%) |
| Prosthetic heart valve | 67 (6.7%) | 5 (3.9%) |
| CAD | 138 (13.8%) | 16 (12.6%) |
| Arrhythmias | 253 (25.3) | 18 (14.2%) |
| A fib/flutter | 150 (15.0%) | 17 (13.4%) |
| VT | 103 (10.3%) | 1 (0.8%) |
| Congenital heart disease | 248 (24.8%) | 4 (3.1%) |
| Hypertension | 188 (18.8%) | 26 (20.5%) |
| Hyperlipidemia | 18 (1.8%) | 8 (6.3%) |
| Diabetes | 96 (9.6%) | 11 (8.7%) |
| CKD | 42 (4.2%) | 6 (4.7%) |
| -CKD | 42 (4.2%) | 6 (4.7%) |
| -ESRD | 0 (0.0%) | 0 (0.0%) |
| OSA | 20 (2.0%) | 2 (1.6%) |
| Pulmonary HTN | 83 (8.3%) | 7 (5.5%) |
| Heart transplant | 51 (5.1%) | 3 (2.4%) |

The model’s efficacy was assessed on the held-out group from the development cohort and the external validation cohort. The performance of the model was assessed utilizing various components of the echocardiogram and clinical information. The first iteration involved the TTE images alone, specifically utilizing videos in the parasternal long axis and apical 4 chamber views, M-mode images, and pulsed wave and continuous wave Doppler images. Then the model was assessed utilizing the structured TTE report (the format of which differed at each site) that included qualitative and quantitative assessments of structure and function, with some demographic information including age, sex, height, weight, and body surface area. The final group (Structured [All]) included all the aforementioned information in addition to concomitant medications, laboratory values, vital signs, and demographics. We assessed combinations of the images and reports as well. We assessed the AI model’s ability to predict peak VO_2_ ≤ 14 mL/kg/min using the data from the echocardiogram (Figure 2). In the development held-out cohort, the model performed well with the area under the receiver operating characteristic curve (AUROC) values ranging from 0.79-0.84. Saliency mapping of PLAX and A4C TTE views indicated greater value in the regions of the left ventricular outflow tract, the medial mitral valve annulus, and the base of the left ventricle (Figure 3). The most influential features involved age, congestion, and inflammation (Figure 4). The best performance was with the inputs of echocardiogram images and the full structured data set (AUROC 0.84, Sensitivity 69.9% given 80% specificity, Specificity 71.5% given 80% sensitivity). The results were consistent in the external validation cohort, with an AUROC of 0.81 (Sensitivity 70.4% given 80% specificity, Specificity 68.2% given 80% sensitivity). There was incremental improvement in the classification metrics as the amount of input increased, with the best regression performance when both the TTE images and the complete structured report were utilized. When the threshold classification value for peak VO2 was adjusted, the model performed consistently well. Utilizing a peak VO2≤12 mL/kg/min (the guideline recommended cutoff for heart transplantation listing), the model performed well for the development held-out cohort (AUROC 0.82) and for the external validation cohort (AUROC 0.74, Table 2). When a more lenient cut-off of VO2≤18 mL/kg/min was applied, the model again demonstrated efficacy in the Columbia held-out (AUROC 0.86) and the external validation cohort (AUROC 0.81). The effectiveness of pretraining the video encoder with Echonet-Dynamic data is shown in Supplemental Table 3.

**Figure 2.**
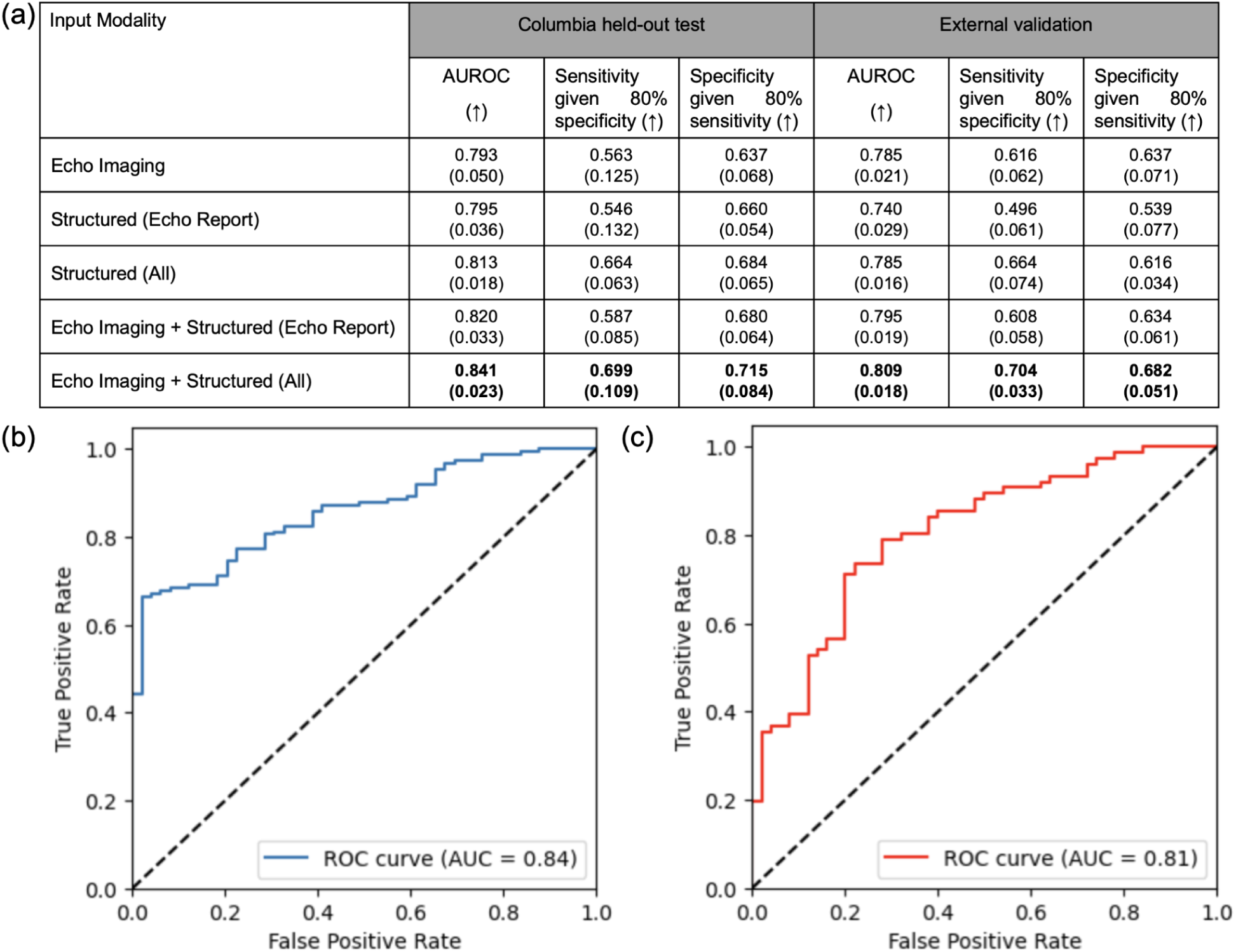
Performance evaluation of the binary classification task for peak VO_2_ (≤14 mL/kg/min). **(a):** Quantitative metrics in the Columbia held-out test data and Cornell data (external validation). Results are averaged across the 5 predefined splits in the Columbia cohort with standard deviations in parentheses. Structured (Echo Report) includes all the structured features in an echo report (echo measurements & findings and some demographics), Structured (All) includes all structured features (echo measurements & findings, vitals, labs, medications, and demographic). AUROC, the area under the receiver operating characteristic curve. **(b) and (c):** The receiver operating characteristic (ROC) curves for (b) Columbia held-out test and (c) WCM & NYP external validation in the first split.

**Figure 3.**
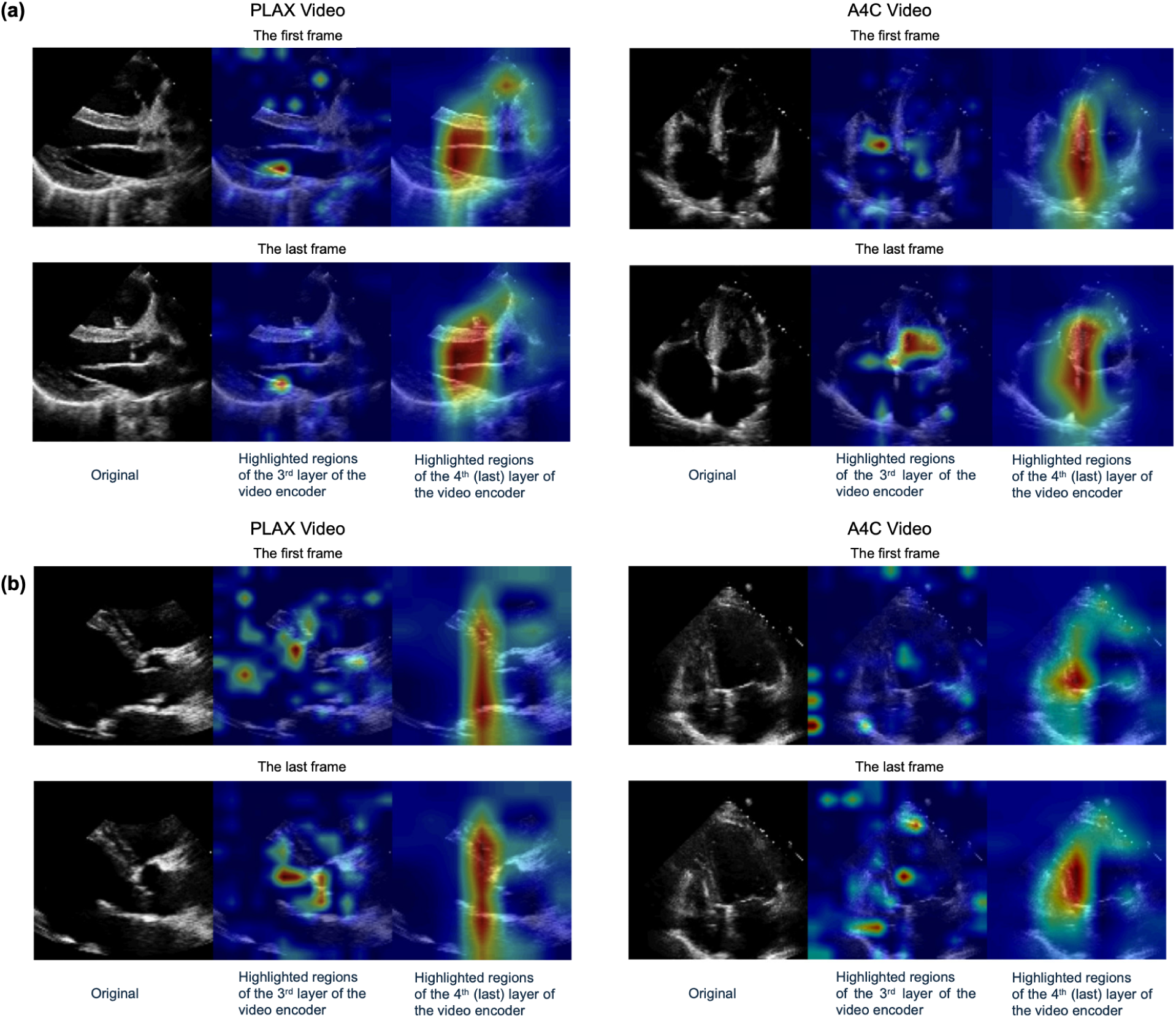
Model interpretability on echo imaging data. Sample visualizations of Grad-CAM saliency maps overlaid on echo videos for two patients: **(a)** peak VO_2_ > 14; **(b)** peak VO_2_ < 14.

**Figure 4.**
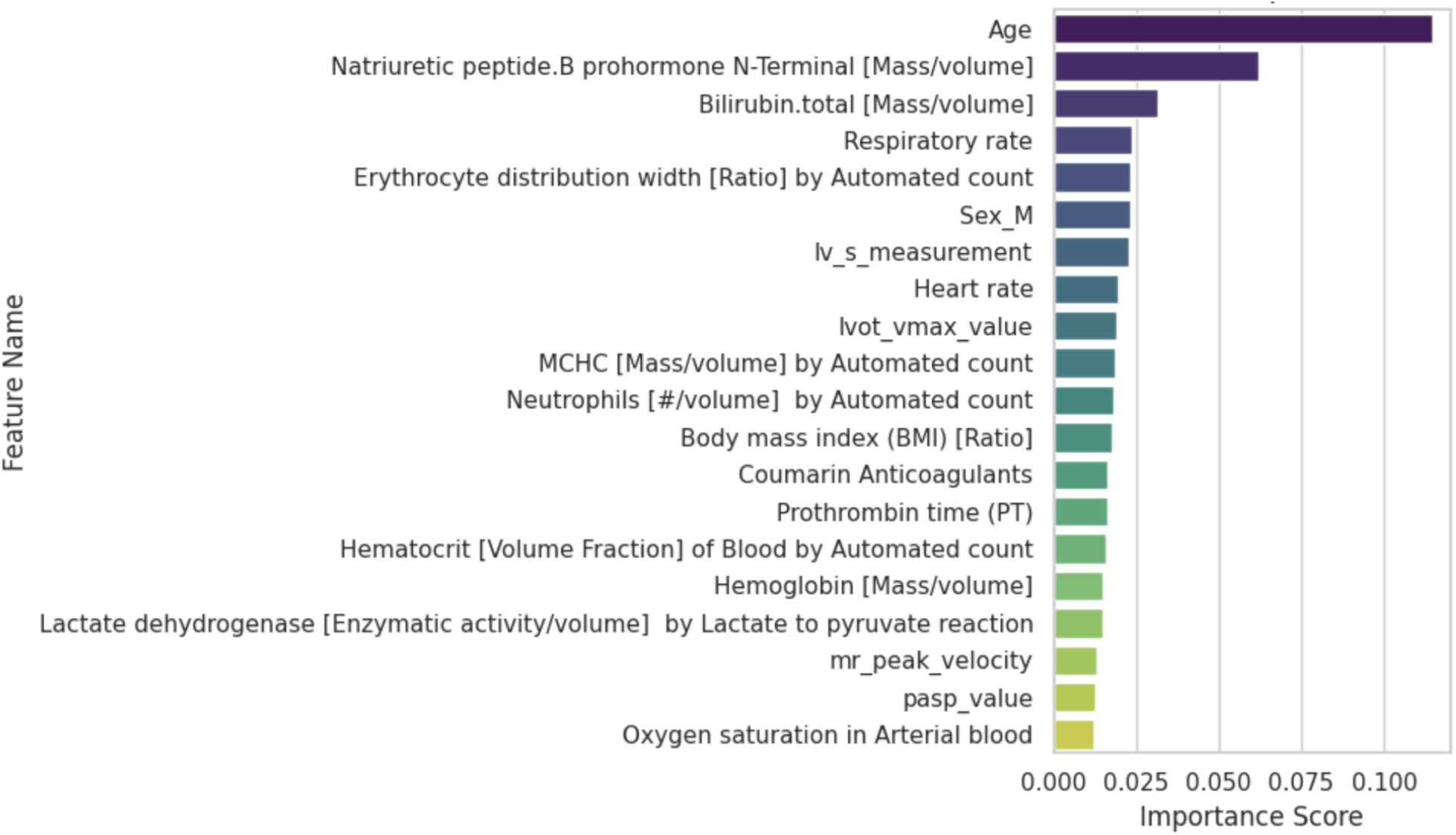
Model interpretability on structured data. The global Shapley value of the top-20 features on the prediction of the model. The features are ranked based on their global Shapley values shown on the x-axis.

**Table 2.** Results of different cutoffs for peak VO_2_ (≤ 12, ≤ 18 mL/kg/min) in the Columbia held-out test data and external validation data. Results are averaged across the 5 predefined splits in the Columbia cohort with standard deviations in parentheses. Structured (Echo Report) includes the structured features in echo report (echo measurements & findings and some demographics), Structured (All) includes all structured features (echo measurements & findings, vitals, labs, medications, and demographic). AUROC, the area under the receiver operating characteristic curve.

| Input Modality | Columbia held-out test |  | External validation |  |
| --- | --- | --- | --- | --- |
| | AUROC<br>($\leq 12$)<br>(†) | AUROC<br>($\leq 18$)<br>(†) | AUROC<br>($\leq 12$)<br>(†) | AUROC<br>($\leq 18$)<br>(†) |
| Echo Imaging | 0.778<br>(0.074) | 0.818<br>(0.037) | 0.701<br>(0.025) | 0.786<br>(0.017) |
| Structured (Echo Report) | 0.745<br>(0.036) | 0.809<br>(0.025) | 0.695<br>(0.018) | 0.726<br>(0.027) |
| Structured (All) | 0.811<br>(0.066) | 0.845<br>(0.035) | 0.744<br>(0.020) | 0.785<br>(0.019) |
| Echo Imaging + Structured<br>(Echo Report) | 0.773<br>(0.046) | 0.830<br>(0.023) | 0.721<br>(0.023) | 0.783<br>(0.008) |
| Echo Imaging + Structured (All) | **0.820**<br>**(0.069)** | **0.856**<br>**(0.032)** | **0.744**<br>**(0.025)** | **0.811**<br>**(0.011)** |

Given the baseline differences between the groups, we assessed the AI model’s consistency stratified by a number of patient baseline demographics. As shown in Table 3, there was consistency between males and females with the AUROC for a peak VO_2_ cutoff of 12 mL/kg/min (0.81 vs. 0.90), 14 mL/kg/min (0.83 vs. 0.89), and 18 mL/kg/min (0.86 vs. 0.89). It was similar for the external validation cohort. There again was consistency for non-white vs. white patients in both the development held-out and external validation cohorts (Table 3). There were differences, however, across age groups. Among patients below the age of 60 the model had good fit and performed well with each peak VO_2_ cutoff for both the internal and external cohorts. The AUROC remained reasonable in the internal validation cohort (>0.72) but declined in the external validation cohort (Table 3). The results further indicate that patients with a shorter interval between cardiopulmonary exercise testing (CPET) and transthoracic echocardiography (TTE) studies—specifically, less than one month—achieved higher AUROC values. This pattern was consistently observed across both internal and external cohorts, highlighting the advantage of utilizing temporally proximate data for predictive modeling in clinical settings. Assessing the cohort stratified by renal function, similar to older age, the model did not perform as well for those with chronic kidney disease (Table 3).

**Table 3.** Results with Echo Imaging + Structured (All) under different subgroups for Columbia held-out test and WCM & NYP external validation. Results are averaged across the 5 predefined splits in the Columbia cohort.

|  | Columbia held-out test |  |  | External validation |  |  |
| --- | --- | --- | --- | --- | --- | --- |
| | AUROC<br>($\leq 14$)(↑) | AUROC<br>($\leq 12$)(↑) | AUROC<br>($\leq 18$)(↑) | AUROC<br>($\leq 14$)(↑) | AUROC<br>($\leq 12$)(↑) | AUROC<br>($\leq 18$)(↑) |
| **Total** | 0.841 | 0.820 | 0.856 | 0.809 | 0.744 | 0.811 |
| **Patients Sex** |  |  |  |  |  |  |
| Male | 0.826 | 0.811 | 0.855 | 0.798 | 0.683 | 0.797 |
| Female | 0.892 | 0.899 | 0.892 | 0.843 | 0.789 | 0.826 |
| **Patients Age** |  |  |  |  |  |  |
| 18 - 59 | 0.881 | 0.875 | 0.868 | 0.857 | 0.827 | 0.784 |
| 60 and above | 0.718 | 0.767 | 0.776 | 0.672 | 0.529 | 0.649 |
| **Race** |  |  |  |  |  |  |
| White | 0.909 | 0.907 | 0.899 | 0.815 | 0.750 | 0.835 |
| Non-White | 0.796 | 0.827 | 0.860 | 0.801 | 0.665 | 0.738 |
| **Days between the Echo and CPET studies** |  |  |  |  |  |  |
| 0 - 30 | 0.870 | 0.860 | 0.903 | 0.844 | 0.803 | 0.812 |
| 30 - 180 | 0.822 | 0.833 | 0.840 | 0.797 | 0.736 | 0.801 |
| **Chronic kidney disease<sup>a</sup>** |  |  |  |  |  |  |
| No | 0.760 | 0.815 | 0.809 | - | - | - |
| Yes | 0.726 | 0.663 | 0.717 | - | - | - |

<sup>a</sup>: Defined by *Glomerular filtration rate/1.73 sq M.predicted [Volume Rate/Area] in Serum, Plasma or Blood by Creatinine-based formula (MDRD) $\leq 60$* . This analysis was conducted only on the Columbia cohort, as the number of patients with CKD in the external validation cohort was insufficient for meaningful evaluation.

## Discussion

Artificial intelligence (in some form) is ubiquitous in our daily lives, however widespread excitement (and disquietude) has abounded in the ChatGPT era. Within medicine, there is similar excitement (and fear) about the role AI can play in patient care moving forward. In this study we sought to leverage echocardiography performed as part of standard clinical care and generate clinically actionable information that previously required additional specialized testing (peak VO_2_). Our principal findings were: 1) A novel AI algorithm demonstrated the ability to predict a patient’s peak VO_2_ across the spectrum of functional impairment (peak VO_2_≤12 mL/kg/min or peak VO_2_≤18 mL/kg/min). 2) The model performed consistently in diverse internal validation and external validation cohorts. 3) The model performance was consistent across gender and race but demonstrated decreased performance in an older population.

The promise of AI to improve medical care is great, and specifically within the field of cardiology tools have been developed to help augment echocardiography. Studies have demonstrated the ability of AI models to accurately quantify ejection fraction, measure chamber size, detect valvular disease, and quantify regurgitation. The automation of these tasks holds the potential to improve the efficiency of the performing sonographer or the interpreting cardiologist, but do not generate novel information. We were able to accomplish that, developing a model that can predict a high risk peak VO_2_ without a concomitant cardiopulmonary exercise test. Utilizing a cut-off of 14 mL/kg/min, guided by the seminal paper by Mancini et al., the model performed well in both internal (AUROC 0.84) and external validation cohorts (AUROC 0.81). We chose 14 mL/kg/min for the screening cut-off as it identifies patients who have or are nearing advanced heart failure, though we felt it was also important to assess the model’s versatility. Utilizing the International Society for Heart and Lung Transplantation guideline recommended cut-off of 12 mL/kg/min (for patients on beta-blockers) for heart transplant listing, the model continued to perform well to identify the sickest patients in internal (AUROC 0.82) and external (0.74) validation cohorts. We also wanted to ensure the model continued to demonstrate efficacy for those with more mild functional limitations, and that was shown with the performance for a peak VO_2_ ≤18 mL/kg/min.

The prevalence of heart failure continues to rise with 6 million (1.7% of the United States population) impacted. There are only about 1,400 advanced heart failure trained physicians in the United States and >40% of United States hospitals with cardiology fellowship programs do not have an advanced heart failure trained physician. Finding who among those 6 million patients should meet one of the 1,400 requires assiduous assessment. Cardiopulmonary exercise testing is a specialized test that can help identify patients who benefit from advanced therapies (e.g. heart transplantation or left ventricular assist device). However, with specialization comes scarcity and CPET is not nearly as ubiquitous as echocardiography. The model developed in this study has the potential to bridge this divide, and with greater precision identify patients who would benefit from referral for more specialized care. We hope that this tool will help democratize care and identify the 97% of patients with advanced heart failure who don’t receive advanced therapies to improve patient outcomes.

The model was not perfect, though, and demonstrated decreased performance in older patients (age > 60 years) and patients with chronic kidney disease. Cardiopulmonary exercise testing is a valid assessment tool in older adults with peak VO_2_ being associated with functional capacity and mortality in this population. However, a reduced peak VO_2_ does not always represent a cardiac deficiency; reduced peak VO_2_ may also be from a pulmonary limitation (perfusion, diffusion, ventilation) or peripheral oxygen extraction limitation (skeletal muscle function, capillary density, mitochondrial function, anemia). It is possible that older patients, like patients with chronic kidney disease, have a greater burden of comorbidities that may impact the peak VO_2_ and not be detected on the echocardiogram. Of note, when the model was trained to predict [the percent of predicted VO2 < 50%] and [VE/VCO2 > 34], it did not perform as well as predicting peak VO2 (Supplemental Table 4). The reduced efficacy in predicting percent predicted VO2 may be because the model already incorporates age and weight into its prediction. Similarly, VE/VCO2 or ventilatory efficiency is not solely attributable to the heart, and can reflect pulmonary hypertension, chronic obstructive pulmonary disease, or other conditions affecting the lungs. Further prospective validation of this model will be important to further assess this observation.

The study has several limitations. First is that the study was a retrospective analysis of patients referred for cardiopulmonary exercise testing, which may introduce confounding by indication; even as the model performed consistently across the spectrum of peak VO_2_. Next, the model was generated utilizing patients from four academic hospitals all in the New York area, potentially limiting generalizability. Additionally, the echocardiograms and the cardiopulmonary exercise tests were contemporaneous but not simultaneous, and it is possible that clinical changes may have occurred between the studies. Finally, we utilized the absolute peak VO_2_ value, while percent predicted peak VO_2_ may also be used for younger patients (<50 years). Nonetheless, the model performed quite well in this age range. Prospective validation will be required.

## CONCLUSIONS

This multimodal AI model effectively categorized patients into low and high risk predicted peak VO_2_ groups using TTE and clinical data. It demonstrates the potential to identify previously unrecognized patients in need of advanced heart failure therapies where CPET is not available.

## Data Availability

The data underlying this article cannot be shared publicly due to restrictions of Weill Cornell Medicine and
Columbia University Irving Medical Center on patient data.

## ABBREVIATIONS LIST

A4C: apical four chamber
AO: aortic valve
CPET: cardiopulmonary exercise testing
CW: continuous wave
LV: left ventricle
MV: mitral valve
PW: pulse wave
RER: respiratory exchange ratio
TTE: transthoracic echocardiography
VO_2_: oxygen consumption

