## Supplementary Information for "Predicting Cardiopulmonary Exercise Testing Performance in Patients Undergoing Transthoracic Echocardiography - An AI Based, Multimodal Model"

### Supplementary Materials

**Supplementary Table 1.** The list of all structured features………...…...……………...………2
**Supplementary Table 2.** The list of echo videos/images to include...………………...………8
**Supplementary Table 3.** Performance evaluation of the binary classification task for peak $VO_2$ ($\leq$14 mL/kg/min) under different settings of pretraining………………………………………9
**Supplementary Table 4.** Performance evaluation of the binary classification task for % Predicted Peak VO2 (<= 50%) and VE/VCO2 (>= 34)...…………………...………………… 10
**Supplementary Figure 1.** Data curation and preprocessing flowchart…...…………...…..……11
**Supplementary Figure 2.** Performance evaluation of the regression task…………...…..……12

Supplemental Tables

**Supplementary Table 1.** The list of all structured features.

| **Category** | **Name** |
|---|---|
| Demo | AGE |
| Demo | SEX |
| Demo | RACE |
| Drug | Loop Diuretics |
| Drug | Coumarin Anticoagulants |
| Drug | Pulmonary Hypertension - Phosphodiesterase Inhibitors |
| Drug | Pulmonary Hypertension - Endothelin Receptor Antagonists |
| Drug | Beta Blockers Non-Selective |
| Drug | Nitrates |
| Drug | Salicylates |
| Drug | Antihypertensive Combinations |
| Drug | Thrombin Inhibitors |
| Drug | Sodium-Glucose Co-Transporter 2 (SGLT2) Inhibitors |
| Drug | Beta Blockers Cardio-Selective |
| Drug | ACE Inhibitors |
| Drug | Alpha-Beta Blockers |
| Drug | Antiarrhythmics Type III |
| Drug | Selective Aldosterone Receptor Antagonists (SARAs) |
| Drug | Cardiovascular Agents Misc. - Combinations |
| Drug | Direct Factor Xa Inhibitors |
| Drug | Antiarrhythmics Type I-B |
| Drug | Intestinal Cholesterol Absorption Inhibitors |
| Drug | Anaphylaxis Therapy Agents |
| Drug | Angiotensin II Receptor Antagonists |
| Drug | Antiadrenergic Antihypertensives |
| Drug | Antianginals-Other |
| Drug | Antiarrhythmics - Misc. |
| Drug | Antiarrhythmics Type I - Nonspecific |
| Drug | Antiarrhythmics Type I-A |
| Drug | Antiarrhythmics Type I-C |
| Drug | Antihyperlipidemics - Combinations |
| Drug | Antihyperlipidemics - Misc. |
| Drug | Bile Acid Sequestrants |
| Drug | Calcium Channel Blockers |
| Drug | Carbonic Anhydrase Inhibitors |

| | |
|---|---|
| Drug | Cardiac Glycosides |
| Drug | Diuretic Combinations |
| Drug | Fibric Acid Derivatives |
| Drug | Gout Agent Combinations |
| Drug | Gout Agents |
| Drug | HMG CoA Reductase Inhibitors |
| Drug | Impotence Agents |
| Drug | Inotropes |
| Drug | Neurogenic Orthostatic Hypotension (NOH) - Agents |
| Drug | Nicotinic Acid Derivatives |
| Drug | Osmotic Diuretics |
| Drug | Peripheral Vasodilators |
| Drug | Platelet Aggregation Inhibitors |
| Drug | Potassium Sparing Diuretics |
| Drug | Proprotein Convertase Subtilisin/Kexin Type 9 Inhibitors |
| Drug | Prostaglandin Vasodilators |
| Drug | Pulmonary Hypertension - Prostacyclin Receptor Agonist |
| Drug | Pulmonary Hypertension - Sol Guanylate Cyclase Stimulator |
| Drug | Sinus Node Inhibitors |
| Drug | Thiazides and Thiazide-Like Diuretics |
| Drug | Transthyretin Stabilizers |
| Drug | Vasoactive Natriuretic Peptides |
| Drug | Vasoactive Soluble Guanylate Cyclase Stimulator (sGC) |
| Drug | Vasodilators |
| Drug | Vasopressors |
| Echo | lv_s_measurement |
| Echo | pasp_less_rap_value |
| Echo | lvot_diameter_value |
| Echo | lvot_area_value |
| Echo | a_wave_velocity |
| Echo | pasp_value |
| Echo | av_peak_gradient |
| Echo | lvpw_measurement |
| Echo | mr_peak_velocity |
| Echo | lv_d_measurement |
| Echo | ivs_measurement |
| Echo | la_area_a2c_value |
| Echo | e_wave_velocity |
| Echo | la_area_a4c_value |

| | |
|---|---|
| Echo | mv_dti_e_prime |
| Echo | mr_alias_velocity |
| Echo | av_vti |
| Echo | lvot_vmean_value |
| Echo | av_mean_gradient |
| Echo | la_volume_index_value |
| Echo | rv_size_value |
| Echo | lvef_value |
| Echo | tr_max_velocity_value |
| Echo | pulmonary_regurgitation_value |
| Echo | e_a_ratio |
| Echo | mv_dti_e_prime_avg |
| Echo | valve_prosthetic_flag |
| Echo | pulmonary_stenosis_value |
| Echo | rap_value |
| Echo | tricuspid_regurgitation_value |
| Echo | mitral_stenosis_value |
| Echo | mr_eroa_pisa |
| Echo | ra_size_value |
| Echo | mitral_valve_prosthetic_flag |
| Echo | lvot_vti_value |
| Echo | la_size_value |
| Echo | mv_dti_e_prime_lateral |
| Echo | mr_volume_flow_rate |
| Echo | mv_dti_a_prime_lateral |
| Echo | mr_pisa_radius |
| Echo | pericardial_effusion_value |
| Echo | mv_dti_e_prime_medial |
| Echo | mitral_regurgitation_value |
| Echo | mr_vti |
| Echo | aortic_stenosis_value |
| Echo | rv_systolic_function_value |
| Echo | lvot_peak_gradient_value |
| Echo | aortic_regurgitation_value |
| Echo | mv_dti_a_prime_medial |
| Echo | lvot_vmax_value |
| Echo | aortic_valve_prosthetic_flag |
| Echo | av_peak_velocity |
| Echo | e_a_ratio_calc_flag |

| | |
|---|---|
| Echo | intra_op_echo_flag |
| Echo | lvad_flag |
| Echo | lvot_mean_gradient_value |
| Echo | mr_fraction_quantitative_doppler |
| Echo | mr_regurg_volume_pisa |
| Echo | mr_volume_quantitative_doppler |
| Echo | mv_dti_a_prime |
| Echo | mv_dti_a_prime_avg |
| Echo | pulmonary_valve_prosthetic_flag |
| Echo | rvad_flag |
| Echo | tricuspid_stenosis_value |
| Echo | tricuspid_valve_prosthetic_flag |
| Echo | unknown_vad_flag |
| Echo | unknown_valve_prosthetic_flag |
| Echo | vad_flag |
| Lab | Natriuretic peptide.B prohormone N-Terminal [Mass/volume] |
| Lab | Erythrocyte distribution width [Ratio] by Automated count |
| Lab | MCHC [Mass/volume] by Automated count |
| Lab | Prothrombin time (PT) |
| Lab | Glomerular filtration rate/1.73 sq M.predicted [Volume Rate/Area] in Serum, Plasma or Blood by Creatinine-based formula (MDRD) |
| Lab | Hematocrit [Volume Fraction] of Blood by Automated count |
| Lab | Calcium [Mass/volume] |
| Lab | Erythrocytes [#/volume] by Automated count |
| Lab | Alkaline phosphatase [Enzymatic activity/volume] |
| Lab | Hemoglobin [Mass/volume] |
| Lab | Bilirubin.total [Mass/volume] |
| Lab | Bilirubin.indirect [Mass/volume] |
| Lab | Glucose [Mass/volume] |
| Lab | Creatinine [Mass/volume] |
| Lab | Neutrophils [#/volume] by Automated count |
| Lab | Neutrophils/100 leukocytes by Automated count |
| Lab | aPTT in Platelet poor plasma by Coagulation assay |
| Lab | Aspartate aminotransferase [Enzymatic activity/volume] |
| Lab | Urea nitrogen [Mass/volume] |
| Lab | Cholesterol.total/Cholesterol in HDL [Mass Ratio] |
| Lab | Protein [Mass/volume] |
| Lab | Phosphate [Mass/volume] |
| Lab | Eosinophils [#/volume] by Automated count |

| | |
|---|---|
| Lab | INR in Platelet poor plasma by Coagulation assay |
| Lab | Lymphocytes [#/volume] by Automated count |
| Lab | MCV [Entitic volume] by Automated count |
| Lab | Potassium [Moles/volume] |
| Lab | Platelet mean volume [Entitic volume] by Automated count |
| Lab | Triglyceride [Mass/volume] |
| Lab | Sodium [Moles/volume] |
| Lab | Monocytes/100 leukocytes by Automated count |
| Lab | Albumin [Mass/volume] |
| Lab | Monocytes [#/volume] by Automated count |
| Lab | Calcium.ionized [Moles/volume] by Ion-selective membrane electrode (ISE) |
| Lab | MCH [Entitic mass] by Automated count |
| Lab | Cholesterol in LDL [Mass/volume] by calculation |
| Lab | Carbon dioxide, total [Moles/volume] |
| Lab | Cholesterol in HDL [Mass/volume] |
| Lab | Alanine aminotransferase [Enzymatic activity/volume] |
| Lab | Leukocytes [#/volume] by Automated count |
| Lab | Hemoglobin [Mass/volume] in Mixed venous blood |
| Lab | Basophils/100 leukocytes by Automated count |
| Lab | Hematocrit [Volume Fraction] of Blood |
| Lab | Platelets [#/volume] by Automated count |
| Lab | Eosinophils/100 leukocytes by Automated count |
| Lab | Lymphocytes/100 leukocytes by Automated count |
| Lab | Immature granulocytes/100 leukocytes |
| Lab | Chloride [Moles/volume] |
| Lab | Magnesium [Mass/volume] |
| Lab | Cholesterol [Mass/volume] |
| Lab | Immature granulocytes [#/volume] |
| Lab | Basophils [#/volume] by Automated count |
| Lab | Nucleated erythrocytes/100 leukocytes [Ratio] by Automated count |
| Lab | Lactate dehydrogenase [Enzymatic activity/volume] by Lactate to pyruvate reaction |
| Lab | pH of Arterial blood |
| Lab | Oxygen [Partial pressure] in Mixed venous blood |
| Lab | Carbon dioxide [Partial pressure] in Arterial blood |
| Lab | Oxygen saturation Calculated from oxygen partial pressure in Arterial blood |
| Lab | Carbon dioxide [Partial pressure] in Mixed venous blood |
| Lab | Bicarbonate [Moles/volume] in Arterial blood |
| Lab | Bilirubin.direct [Mass/volume] |

| | |
|---|---|
| Lab | Oxygen [Partial pressure] in Arterial blood |
| Lab | Bicarbonate [Moles/volume] in Mixed venous blood |
| Lab | Hemoglobin [Mass/volume] in Arterial blood |
| Lab | pH of Mixed venous blood |
| Lab | Base excess in Arterial blood by calculation |
| Lab | Base excess in Mixed venous blood by calculation |
| Lab | Calcium.ionized [Moles/volume] |
| Lab | Carbon dioxide, total [Moles/volume] in Arterial blood |
| Lab | Carbon dioxide, total [Moles/volume] in Mixed venous blood |
| Lab | Lactate [Moles/volume] |
| Lab | Nucleated erythrocytes [#/volume] by Automated count |
| Lab | Oxygen saturation in Mixed venous blood |
| Lab | Platelets reticulated/100 platelets by Automated count |
| Vital | Body mass index (BMI) [Ratio] |
| Vital | Heart rate |
| Vital | Systolic blood pressure |
| Vital | Respiratory rate |
| Vital | Body surface area |
| Vital | Oxygen saturation in Arterial blood |
| Vital | Body temperature |
| Vital | Respiration rhythm |
| Vital | Diastolic blood pressure |
| Vital | Pulse |
| Vital | Oxygen saturation in Arterial blood by Pulse oximetry |

**Supplementary Table 2.** The list of echo videos/images to include.

| **Category** | **View to Include** |
|---|---|
| Video | Apical 4 Chamber (A4C) |
| Video | Parasternal Long Axis (PLAX) |
| M-Mode Image | Parasternal Long Axis - Aorta (PLAX-AO) |
| M-Mode Image | Parasternal Long Axis - Mitral Valve (PLAX-MV) |
| M-Mode Image | Parasternal Long Axis - Aorta Valve (PLAX-AV) |
| Doppler Image | Spectral Doppler: CW - TR Vmax |
| Doppler Image | Spectral Doppler: CW - PV peak gradient |
| Doppler Image | Spectral Doppler: PW - MV peak E Vel |

**Supplementary Table 3. Performance evaluation of the binary classification task for peak $VO_2$ (≤14 mL/kg/min) under different settings of pretraining.** Results are averaged across the 5 predefined splits in the Columbia cohort with standard deviations in parentheses.

| Input Modality | Pretrained Video Encoder | Columbia held-out test | | | External validation | | |
|---|---|---|---|---|---|---|---|
| | | AUROC (↑) | Sensitivity given 80% specificity (↑) | Specificity given 80% sensitivity (↑) | AUROC (↑) | Sensitivity given 80% specificity (↑) | Specificity given 80% sensitivity (↑) |
| Echo Imaging | No | 0.775 (0.054) | 0.609 (0.095) | 0.519 (0.133) | 0.698 (0.026) | 0.443 (0.089) | 0.440 (0.024) |
| Echo Imaging | Yes | 0.793 (0.050) | 0.563 (0.125) | 0.637 (0.068) | 0.785 (0.016) | 0.616 (0.062) | 0.637 (0.071) |
| Echo Imaging + Structured (All) | No | 0.836 (0.024) | 0.666 (0.113) | 0.691 (0.094) | 0.797 (0.009) | 0.581 (0.050) | 0.678 (0.060) |
| Echo Imaging + Structured (All) | Yes | **0.841 (0.023)** | **0.699 (0.109)** | **0.715 (0.084)** | **0.809 (0.018)** | **0.704 (0.033)** | **0.682 (0.051)** |

**Supplementary Table 4. Performance evaluation of the binary classification task for % Predicted Peak VO2 (<= 50%) and VE/VCO2 (>= 34).** Results are averaged across the 5 predefined splits in the Columbia cohort with standard deviations in parentheses.

| Input Modality | Columbia held-out test | |
|---|---|---|
| | % Predicted Peak VO2 <= 50% AUROC(↑) | VE/VCO2 >= 34 AUROC (↑) |
| Echo Imaging | 0.745 (0.028) | 0.765 (0.029) |
| Structured (All) | 0.786 (0.063) | 0.779 (0.036) |
| Echo Imaging + Structured (All) | **0.806 (0.043)** | **0.816 (0.025)** |

### Supplemental Figures

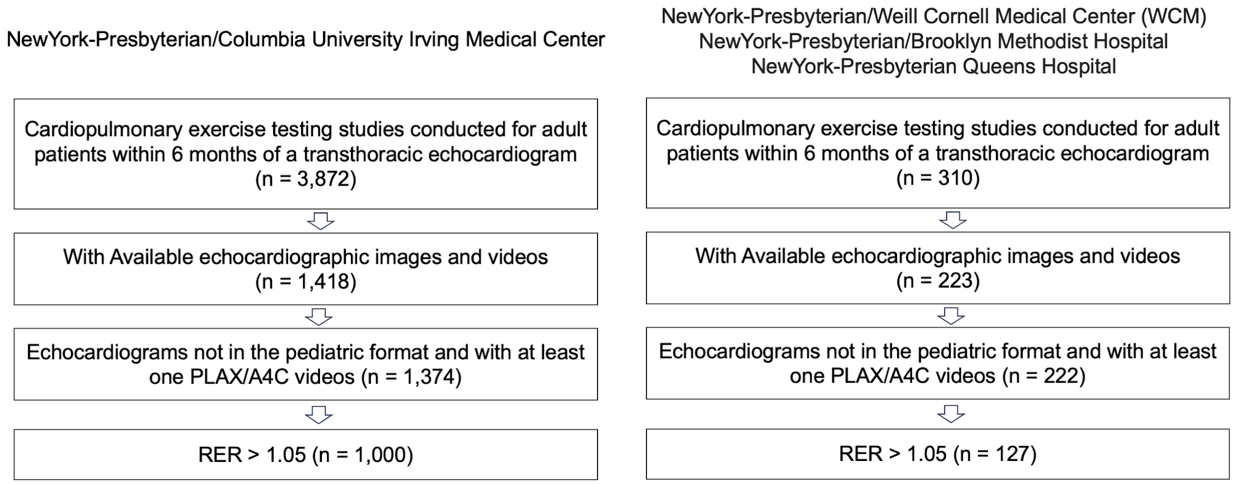


**Supplementary Figure 1.** Data curation and preprocessing flowchart.

(a)

| Input Modality | Columbia held-out test | | | External validation | | |
|---|---|---|---|---|---|---|
| | MAE(↓) | RMSE(↓) | R2(↑) | MAE(↓) | RMSE(↓) | R2(↑) |
| Echo Imaging | 4.43 (0.17) | 5.73 (0.38) | 0.446 (0.036) | 5.70 (0.14) | 7.08 (0.14) | 0.320 (0.026) |
| Structured (Echo Report) | 4.35 (0.26) | 5.79 (0.44) | 0.432 (0.080) | 5.43 (0.34) | 6.89 (0.39) | 0.354 (0.073) |
| Structured (All) | 3.98 (0.27) | 5.21 (0.48) | 0.542 (0.055) | **5.11 (0.08)** | 6.48 (0.09) | 0.431 (0.016) |
| Echo Imaging + Structured (Echo Report) | 4.14 (0.26) | 5.41 (0.50) | 0.506 (0.066) | 5.39 (0.26) | 6.77 (0.29) | 0.378 (0.053) |
| Echo Imaging + Structured (All) | **3.89 (0.28)** | **5.10 (0.50)** | **0.560 (0.057)** | 5.17 (0.08) | **6.46 (0.07)** | **0.434 (0.012)** |

(b)

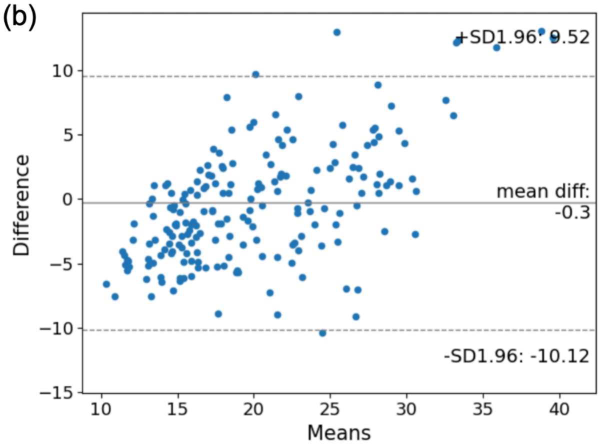


(c)

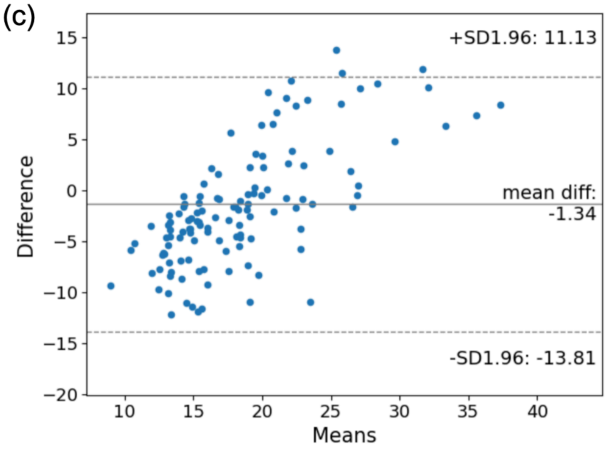


**Supplementary Figure 2. Performance evaluation of the regression task for peak VO2 (mL/kg/min). (a)**: Quantitative metrics in the Columbia held-out test data and Cornell data (external validation). Results are averaged across the 5 predefined splits in the Columbia cohort with standard deviations in parentheses. *Structured (Echo Report)* includes the structured features in echo report (echo measurements & findings and some demographics), *Structured (All)* includes all structured features (echo measurements & findings, vitals, labs, medications, and demographic). MAE, mean absolute error; RMSE, Root mean square deviation. **(b) and (c)**: the Bland-Altman plot of predicted peak $VO_2$ compared with its ground-truth value for (b) Columbia held-out test and (c) external validation in the first split. In each subfigure, the x-axis represents the averaged value of predicted and ground-truth peak $VO_2$, while the y-axis represents the difference between predicted and ground-truth peak $VO_2$.
